# Measuring Subjective Cognitive Decline in Older Adults: Harmonization between the Cognitive Change Index and the Measurement of Everyday Cognition Instruments

**DOI:** 10.1101/2021.09.27.21262840

**Authors:** Lindsey F Wells, Shannon L Risacher, Brenna C McDonald, Martin R Farlow, Jared Brosch, Sujuan Gao, Liana G Apostolova, Andrew J Saykin, for the Alzheimer’s Disease Neuroimaging Initiative

**Author notes:** Data used in preparation of this article were obtained from the Alzheimer’s Disease Neuroimaging Initiative (ADNI) database (adni.loni.usc.edu). As such, the investigators within the ADNI contributed to the design and implementation of ADNI and/or provided data but did not participate in analysis or writing of this report. A complete listing of ADNI investigators can be found at: http://adni.loni.usc.edu/wp-content/uploads/how_to_apply/ADNI_Acknowledgement_List.pdf. Corresponding Author:* Dr. Andrew J. Saykin, 355 W. 16th St., Suite 4100, Center for Neuroimaging, Department of Radiology and Imaging Sciences, Indiana University School of Medicine, Indianapolis, Indiana 46202, USA.

## Abstract

**Background:** Self and informant (proxy or study partner) reports of everyday cognitive functioning have been shown to be associated with incipient neurodegenerative disease. The 20-item Cognitive Change Index (CCI) and the 39-item Measurement of Everyday Cognition (ECog) were each developed to characterize early subjective changes in cognitive function.

**Objective:** We examined the relationship between CCI and ECog self and informant evaluations to determine content overlap and provide a co-calibration for converting between these widely used instruments.

**Methods:** 950 participants (57.1% female, mean age=71.2yrs) from ADNI and the Indiana ADRC with self evaluations and 279 participants (60.9% female, mean age=71.8yrs) with informant evaluations (Indiana ADRC) were included. Analyzed variables for the CCI and ECog included domain mean scores, memory domain total scores, and total scores for all items. Pearson correlations, regression analyses, and frequency distributions were used to assess the relationship between CCI and ECog. Sex, years of education, race, *APOE* ε4 carrier status, and baseline diagnosis were also analyzed as potentially relevant covariates.

**Results:** CCI and ECog total scores were highly correlated for the self (r=0.790, p<0.001) and informant (r=0.860, p<0.001) versions. Frequency distributions of total scores were generated and self and informant histograms were plotted separately. Quadratic regressions for self (r^2^=0.682) and informant (r^2^=0.863) scores were used to create a translation table between the CCI and ECog total scores.

**Conclusion:** Self and informant total scores can be harmonized and translated between the CCI and ECog to facilitate cross-study and longitudinal assessment of perceived cognitive change, an important patient-reported outcome.

## INTRODUCTION

Prior to the onset of objective cognitive impairment, individuals with preclinical Alzheimer’s disease (AD) often present with self report of cognitive decline or reports of cognitive decline by someone who knows them well [1]. Subjective cognitive decline (SCD) is a term given to the period of preclinical dementia during which some decline in everyday cognitive functioning is subjectively recognized by an individual, but there is no evidence of a deficit on objective cognitive tests [2,3]. The presence of significant subjective cognitive concerns can serve as an additional factor to identify individuals at increased risk of progressing to mild cognitive impairment (MCI) or dementia. In previous studies, SCD has been shown to be associated with relevant AD biomarker abnormalities [4-6] and later a diagnosis of MCI or AD [1]. These concerns are important for early detection of dementia because they have been shown to present as early as 15 years prior to diagnosis of MCI or AD [1].

A variety of cognitive assessment tools are available to screen for the early stages of neurodegenerative disease. The Mini-Mental State Exam [7] and Montreal Cognitive Assessment [8] are objective cognitive assessments commonly used for dementia screening; however, they have limitations in terms of detecting these early stages [9]. The 20-item Cognitive Change Index (CCI) [10] and the 39-item Measurement of Everyday Cognition (ECog) [9] are two tools developed specifically to assess SCD. The 12-item CCI-12 is a shortened version, consisting of only the memory domain, that has also been used to evaluate SCD. Both tools have subject (self) and informant (proxy or study partner) versions to evaluate the extent of cognitive concerns for memory, executive functioning, and language. The ECog has additional items to assess the visual-spatial domain. The CCI and ECog each have a different Likert scale and total number of items. There is currently no method for directly comparing or translating between CCI and ECog scores.

The aim of this study is to define the relationship between these tools to improve longitudinal assessment of cognitive concerns. SCD has been shown to be associated with a higher likelihood of onset of neurodegenerative disease [11, 12], which is important, as it informs clinicians about possible risk factors. This cross-sectional study co-calibrates and provides a means to harmonize and translate data between the CCI and the ECog to promote future cross-study analyses.

## MATERIALS AND METHODS

### Participants

Data used in the preparation of this article were obtained from the Alzheimer’s Disease Neuroimaging Initiative (ADNI) and the Indiana Alzheimer’s Disease Research Center (IADRC) including participants from the Indiana Memory and Aging Studies (IMAS) phase and IADRC phases.

ADNI was launched in 2003 by the National Institute on Aging (NIA), the National Institute of Biomedical Imaging and Bioengineering (NIBIB), the Food and Drug Administration (FDA), and private pharmaceutical companies and non-profit organizations. One of the primary goals of ADNI has been to test whether neuropsychological assessment can be used to measure the progression of mild cognitive impairment (MCI) and early Alzheimer’s disease (AD). See the ADNI website (http://adni.loni.usc.edu) for more details. The IADRC, including the IMAS and IADRC phases, likewise includes objective and subjective evaluation of cognitive function as part of longitudinal observational studies of older adults at risk for and with clinical AD. Informed consent for all included data was obtained by the respective study protocol according to the Declaration of Helsinki.

Data were acquired between 2012 and 2019, and included 639 participants from ADNI, 326 participants from IADRC, and 20 participants from the initial phase of IMAS. At baseline, 615 participants were classified as cognitively normal (CN), 261 were considered to have MCI, and 109 were diagnosed with AD. Participants were included if they or their informant had completed equal to or more than 75% of the items on both the CCI and the ECog, and if the assessments were completed less than a year apart. In the ADNI protocol, the CCI is obtained during the screening visit, whereas the ECog is administered at baseline visit. On average, the difference in time was less than 2 months. In the IMAS and IADRC protocols, the assessments can be completed beforehand or at the time of visit, but only the visit date was recorded. 950 self reports and 279 informant reports were available for analysis. Sex, years of education, race, *APOE* ε4 carrier status, baseline diagnosis, and time difference between administration of the assessments were analyzed as potentially relevant covariates. This study was approved by the Indiana University (IU) Institutional Review Board for data access and analysis.

Clinical diagnosis of CN, MCI, or AD was determined based on the diagnostic criteria [13]. CN subjects were defined as having no cognitive concerns and preserved functional status [13]. Subjects with MCI were defined as having mild objective cognitive impairment with preserved functional status [13]. Subjects with AD were defined based on moderate to severe cognitive impairment and low functional status [13]. The ADNI database includes participants ages 55 to 90. Participants from IADRC and IMAS phases were included if their age was greater than 50. Participants were not included if they had neurological or psychiatric disease such as major depressive disorder, bipolar disorder or schizophrenia other than AD. Notably, minor depression and anxiety were not excluded.

### Cognitive Change Index (CCI) and Measurement of Everyday Cognition (ECog)

The CCI consists of 20 items that ask participants and their informants to rate the participant’s cognitive status relative to the previous 5 years [10]. The self and informant evaluation forms are available from the authors (contact Dr. Saykin at). Each item is rated on a Likert scale out of 5 possible points (1 = no change or better, 2 = minimal change or slight/occasional problem, 3 = some change or mild problem, 4 = clearly noticeable change or moderate problem, 5 = much worse or severe problem). There are 12 items in the episodic memory domain, 5 items in the executive functioning domain, and 3 items in the language domain. The CCI-12 consists of the memory domain only and has also been used to define SCD.

The ECog includes 39 items that ask participants and their informants to rate the participant’s cognitive status relative to 10 years ago (contact Dr. Tomaszewski Farias at for a copy of the 39 item ECog) [9]. Each item is rated out of 4 possible points (1 = better or no change, 2 = questionable/occasionally worse, 3 = consistently a little worse, 4 = consistently much worse, 9 = don’t know). There are 8 items in the memory domain, 9 items in the language domain, 15 items in the executive function domain (in three sub-domains: planning, organization, and divided attention), and 7 items in the visual-spatial domain. Of note, the CCI does not have a visual-spatial domain.

For both the CCI and ECog a mean score for each domain, a total score, and a total memory score were calculated to determine which calculation had the highest correlation between the assessment tools. Domain mean scores were calculated by dividing the sum of each domain by the number of items. Total scores were determined by adding together the points for each item in the assessment. Total memory scores were calculated by adding the points for each item in the memory domain alone.

### Statistical analysis

Descriptive statistics of participant characteristics were calculated including frequency for categorical variables and mean and standard deviation for continuous measures. Characteristics including age, sex, years of education, race, *APOE* ε4 carrier status, CCI scores, and ECog scores were compared between ADNI, IADC, and IMAS self and informant reports. Since participants were included if they had completed at least 75% of both assessments, there was some item level missing data. Linear interpolation was performed using SPSS 27 to impute this missing data. One-way ANOVA was used to compare total scores for each measure between diagnostic subgroups. Associations between CCI and ECog mean domain scores for the memory, executive function, and language domains were assessed using Pearson correlation in the self and informant assessments. Associations between CCI and ECog total scores for the memory domain were also assessed using Pearson correlation in the self and informant assessments. Frequency distributions of self and informant CCI and ECog total scores were plotted using a bin width of 10 and a bin center starting at 20 and 40 for the CCI and ECog respectively. Polynomial regression analyses of CCI versus ECog total scores were determined separately for the self and informant assessments. Quadratic regression provided the greatest adjusted R-square value compared to linear and cubic regressions. Therefore, a quadratic equation was used to generate a crosswalk table [14] between the CCI and ECog. Statistical analyses were performed using SPSS 27, and plots were generated using GraphPad Prism 9.

## RESULTS

### Participant demographic characteristics

There were 639 participants from the ADNI cohort and 311 participants from the IADRC study and IMAS studies in the self-report group. Demographic characteristics are shown in Table 1. The mean age of all participants in the self-report group was 71.2 years (range: 52-93 years), with 57.1% female and 83.6% Non-Hispanic White. The mean education for participants in the self-report group was 16.4 years (range: 7-23). The mean self-report total score using the imputed data was 60.7 +/- 20.2 (39-144) for the ECog and 37.3 +/- 14.5 (20-88) for the CCI.

**Table 1.** Demographic Characteristics of the Cohorts

Demographic Characteristics of the Cohorts
| A) Demographic Characteristics of Self-Assessment Cohort |  |  |  |  |  |
| --- | --- | --- | --- | --- | --- |
|  | CN (n=612) | MCI (n=255) | AD (n=83) | p-value | Pairwise* |
| Age (years) | 70.6 (6.8) | 72.1 (7.6) | 73.1 (9.3) | 0.001 | MCI, AD>CN |
| Education (years) | 16.5 (2.5) | 16.2 (2.6) | 16.0 (2.3) | 0.06 | None |
| Gender (M, F) | 228, 384 | 137, 118 | 43, 40 | <0.001 | n/a |
| Race/Ethnicity (%NHW) | 85.1% | 80.4% | 82.0% | 0.53 | n/a |
| APOE Genotype (% ε4+)<sup>1</sup> | 34.1% | 48.7% | 60.0% | <0.001 | n/a |
| CDR-Sum of Boxes<sup>2</sup> | 0.1 (0.3) | 1.5 (1.0) | 4.4 (1.9) | <0.001 | AD>MCI>CN |
| NPI-Q Total<sup>3</sup> | 1.1 (2.6) | 4.2 (6.0) | 7.9 (8.5) | <0.001 | AD>MCI>CN |
| GDS Total<sup>4</sup> | 1.0 (1.3) | 2.1 (2.0) | 1.9 (1.8) | <0.001 | AD, MCI>CN |
| MoCA Total Score<sup>5</sup> | 26.0 (2.7) | 22.6 (3.2) | 17.2 (4.7) | <0.001 | CN>MCI>AD |
| Story Immediate Recall (z)<sup>6</sup> | 0.2 (0.9) | -1.2 (1.1) | -2.6 (0.8) | <0.001 | CN>MCI>AD |
| Story Delayed Recall (z)<sup>6</sup> | 0.2 (0.9) | -1.1 (1.0) | -2.4 (0.7) | <0.001 | CN>MCI>AD |
| List Learning Immediate (z)<sup>7</sup> | -0.1 (1.0) | -1.1 (1.0) | -2.4 (0.8) | <0.001 | CN>MCI>AD |
| List Learning Delayed (z)<sup>7</sup> | -0.2 (1.2) | -1.4 (1.3) | -2.5 (0.7) | <0.001 | CN>MCI>AD |
| Naming Score (z)<sup>8</sup> | -1.0 (3.0) | -0.8 (2.2) | -2.5 (3.4) | <0.001 | CN, MCI>AD |
| <sup>1</sup> Missing 62 participants (23 CN, 31 MCI, 8 AD) |  | <sup>4</sup> Missing 1 participant (1 MCI) |  | <sup>7</sup> Missing 33 participants (20 CN, 10 MCI, 2 AD) |  |
| <sup>2</sup> Missing 3 participants (2 CN, 1 MCI) |  | <sup>5</sup> Missing 16 participants (5 CN, 7 MCI, 4 AD) |  | <sup>8</sup> Missing 7 participants (4 CN, 2 MCI, 1 AD) |  |
| <sup>3</sup> Missing 22 participants (14 CN, 8 MCI) |  | <sup>6</sup> Missing 4 participants (3 CN, 1 MCI) |  | * Bonferroni corrected p<0.05 |  |
| B) Demographic Characteristics of Participants in Informant-Assessment Cohort |  |  |  |  |  |
|  | CN (n=163) | MCI (n=75) | AD (n=41) | p-value | Pairwise* |
| Age (years) | 71.0 (8.3) | 73.0 (7.7) | 72.6 (11.6) | 0.20 | None |
| Education (years) | 15.9 (2.7) | 16.0 (2.8) | 15.3 (3.3) | 0.44 | None |
| Gender (M, F) | 63, 100 | 31, 44 | 15, 26 | 0.87 | n/a |
| Race/Ethnicity (%NHW) | 88.3% | 74.7% | 75.6% | 0.02 | n/a |
| APOE Genotype (% ε4+)<sup>9</sup> | 33.3% | 61.3% | 61.0% | <0.001 | n/a |
| CDR-Sum of Boxes<sup>10</sup> | 0.2 (0.5) | 1.4 (1.0) | 6.1 (4.6) | <0.001 | AD>MCI>CN |
| NPI-Q Total<sup>11</sup> | 1.6 (2.5) | 3.6 (3.8) | 7.2 (6.2) | <0.001 | AD>MCI>CN |
| GDS Total<sup>12</sup> | 1.4 (1.7) | 3.0 (2.6) | 2.3 (2.5) | <0.001 | AD, MCI>CN |
| MoCA Total Score<sup>13</sup> | 26.0 (2.9) | 21.6 (3.3) | 14.9 (6.1) | <0.001 | CN>MCI>AD |
| Story Immediate Recall (z)<sup>14</sup> | 0.3 (1.0) | -1.2 (1.2) | -2.5 (1.0) | <0.001 | CN>MCI>AD |
| Story Delayed Recall (z)<sup>14</sup> | 0.2 (1.0) | -1.0 (1.3) | -2.1 (1.1) | <0.001 | CN>MCI>AD |
| List Learning Immediate (z)<sup>15</sup> | -0.1 (1.0) | -1.4 (0.9) | -2.4 (0.9) | <0.001 | CN>MCI>AD |
| List Learning Delayed (z)<sup>15</sup> | 0.1 (1.0) | -1.6 (1.0) | -2.5 (0.8) | <0.001 | CN>MCI, AD |
| Naming Score (z)<sup>16</sup> | -0.4 (2.3) | -1.1 (2.3) | -1.8 (2.7) | 0.001 | CN, MCI>AD |
| <sup>9</sup> Missing 1 participant (1 CN) |  | <sup>12</sup> Missing 3 participants (3 AD) |  | <sup>15</sup> Missing 32 participants (10 CN, 7 MCI, 15 AD) |  |
| <sup>10</sup> Missing 1 participant (1 CN) |  | <sup>13</sup> Missing 5 participants (2 MCI, 3 AD) |  | <sup>16</sup> Missing 5 participants (5 AD) |  |
| <sup>11</sup> Missing 4 participants (4 CN) |  | <sup>14</sup> Missing 5 participants (5 AD) |  | * Bonferroni corrected p<0.05 |  |

There were 279 participants from the IADRC and IMAS studies in the informant-report group. 244 of these reports were for individuals for whom self-report data was also included. The mean age of all participants in the informant-report group was 71.8 years (range: 52-95 years), with 60.9% female and 82.8% Non-Hispanic White. The mean education for participants in the informant-report group was 15.9 years (range: 4-22). The mean informant-based total score for the ECog was 66.5 +/- 30.7 (range: 39-156) and for the CCI was 39.5 +/- 21.0 (range: 20-100).

### Mean scores in each diagnostic subgroup

Participants from the self versus informant assessments had significantly different scores when stratified by diagnostic subgroup (Table 2). The CCI and ECog total scores were the lowest in the CN group as anticipated, with intermediate scores in MCI, and the highest scores in AD.

**Table 2.** Total scores of ECog (39 items) and CCI (20 items) by diagnostic subgroup

|  |  | **CN** | **MCI** | **AD** | **p-value** |
| --- | --- | --- | --- | --- | --- |
| **SELF** | **n** | 612 | 255 | 83 |  |
|  | **ECog** | 54.1 ± 14.1 | 71.9 ± 23.3 | 75.3 ± 24.6 | <0.01** between CN-AD and CN-MCI |
|  | **CCI-20** | 31.8 ± 10.4 | 46.9 ± 15.7 | 48.4 ± 14.6 | <0.01** between CN-AD and CN-MCI |
| **INFORMANT** | **n** | 163 | 75 | 41 |  |
|  | **ECog** | 50.5 ± 16.0 | 76.6 ± 25.9 | 111.7 ± 30.9 | <0.01** between all groups |
|  | **CCI-20** | 27.4 ± 10.2 | 47.3 ± 15.2 | 73.7 ± 17.8 | <0.01** between all groups |

CN=Cognitively Normal; MCI=Mild Cognitive Impairment; AD=Alzheimer's Disease
\*\*p<0.01

### Associations between CCI and ECog scores

Pearson correlations (Table 3) showed that the CCI and ECog were significantly associated (range r=0.610 to 0.860, all p < 0.001). The highest correlation for the self and the informant assessments was between the total scores.

**Table 3.** Pearson correlations between the CCI and ECog paired scores

|  | **Domain** | **PEARSON CORRELATIONS** |  |
| --- | --- | --- | --- |
|  |  | **Self**<br>**n = 950** | **Informant**<br>**n = 279** |
| **Mean Domain Score** | Memory | 0.758** | 0.856** |
|  | Language | 0.650** | 0.774** |
|  | Executive function | 0.610** | 0.741** |
|  | All domains | 0.785** | 0.860** |
| **Total Score** | Memory | 0.759** | 0.858** |
|  | All domains | 0.790** | 0.860** |

\*\*p<0.01

We evaluated frequency distributions of the 950 self reports and the 279 informant reports total scores (Figure 1). Based on the histogram of self reports, the highest number of entries fell between a score of 25 to 35 for the CCI and 45 to 55 for the ECog. For the informant, the highest number of entries fell between 20 to 25 for the CCI and 39 to 45 for the ECog. The difference between the CCI and ECog is explained by the 19-point difference in minimum total score. Quadratic regression analysis (Figure 2) showed the best fit between the CCI and ECog total scores. All diagnostic subgroups were included in the analysis. A quadratic regression equation was generated to create a conversion table between the CCI-20 and the ECog-39. Separate tables were made for the self-report (Table 4) and informant-report (Table 5) versions for the. Similar tables were made to translate between the CCI-12 and the ECog (Table 6 and Table 7).

**Figure 1.**
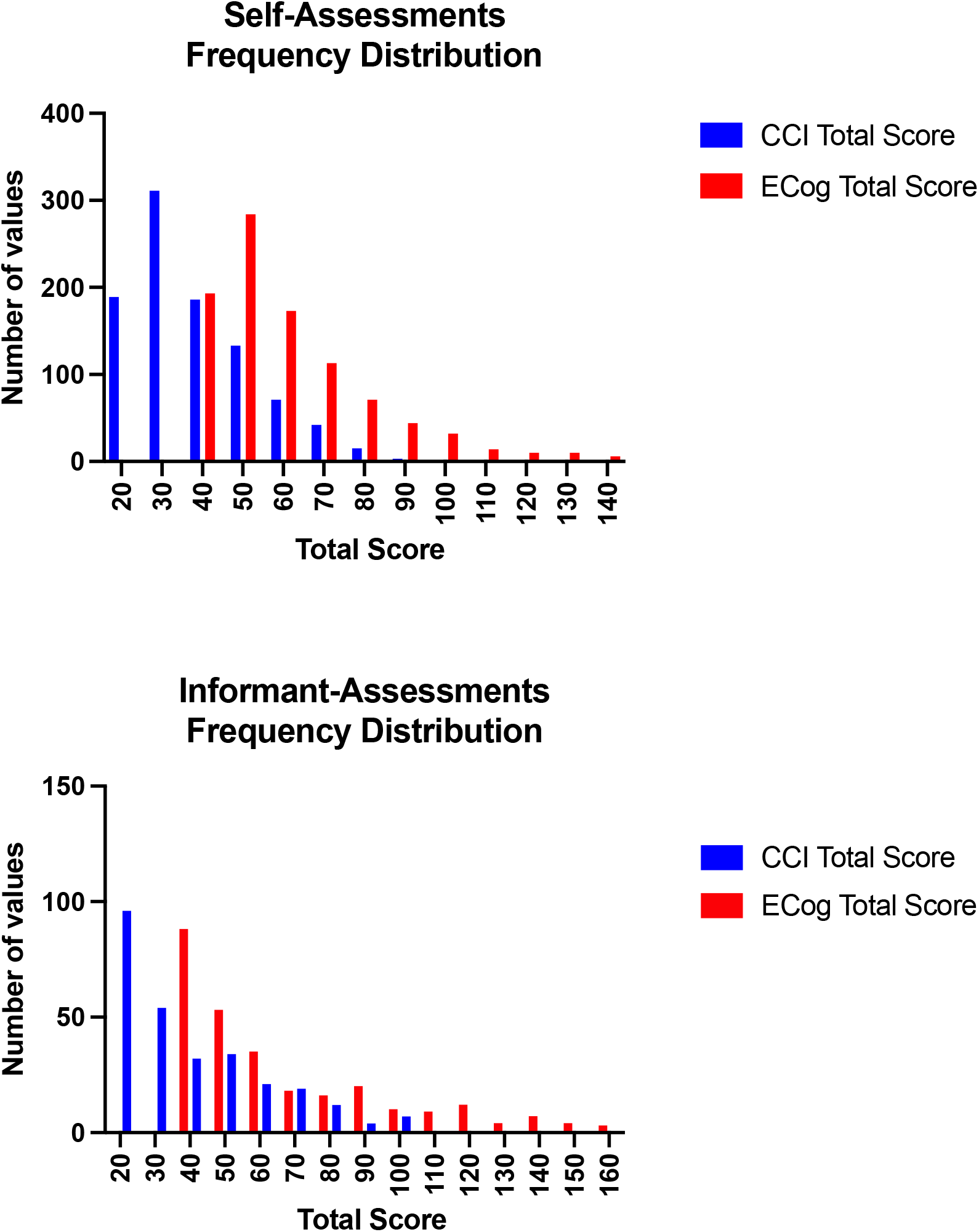
Frequency distributions showing the patterns between CCI 20-item and ECog 39-item total scores in the self and informant assessments. CCI, Cognitive Change Index; ECog, Measurement of Everyday Cognition

**Figure 2.**
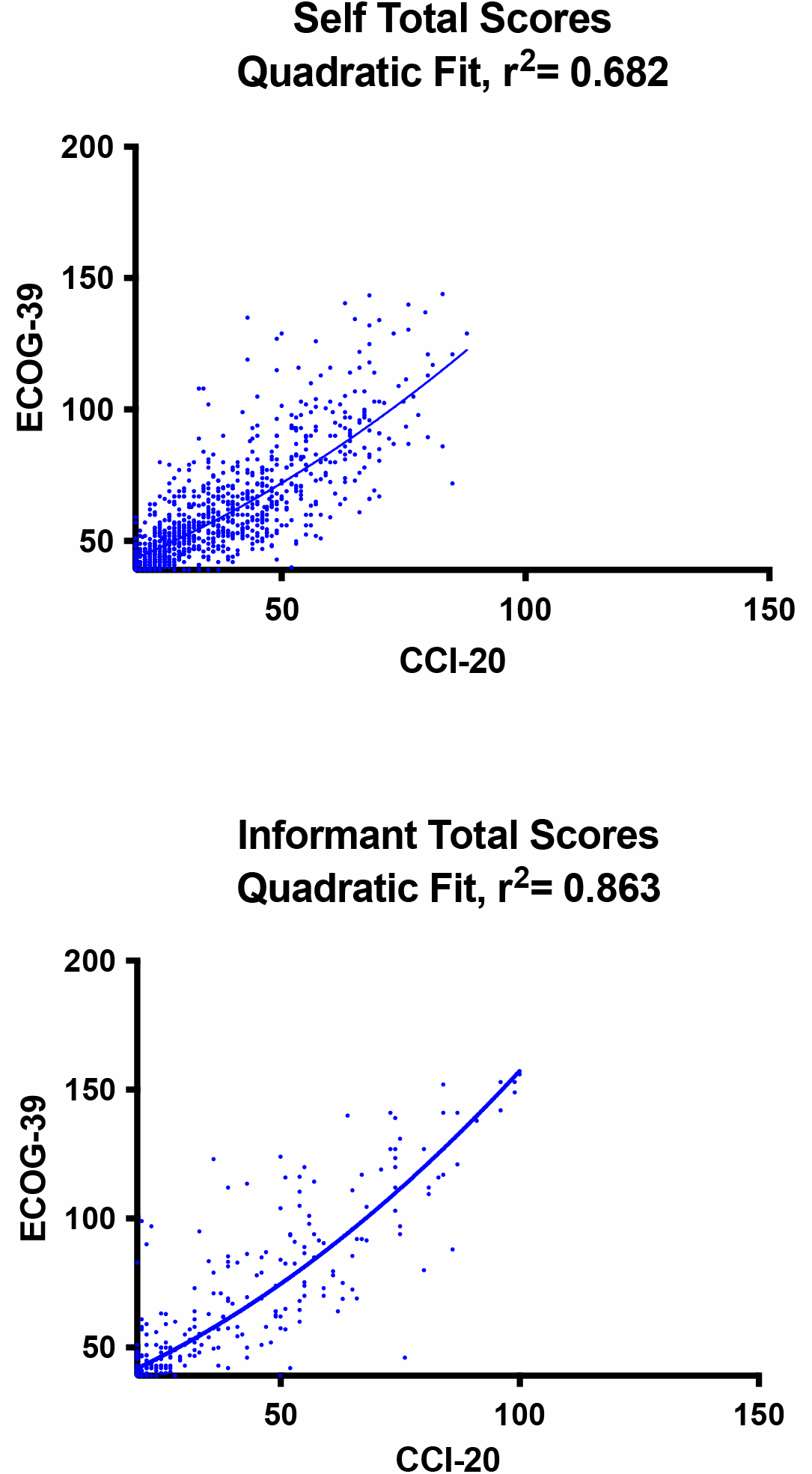
Quadratic regression showing the relationship between CCI 20-item and ECog 39-item total scores in the self and informant assessments. CCI, Cognitive Change Index; ECog, Measurement of Everyday Cognition

**Table 4.** CCI (20 items) self-assessment mapped to ECog (39 items) self-assessment using Quadratic Regression

| **CCI-20** | **ECog** | **CCI-20** | **ECog** | **CCI-20** | **ECog** | **CCI-20** | **ECog** |
| --- | --- | --- | --- | --- | --- | --- | --- |
| **20** | **43** | 41 | 62 | **62** | **86** | 83 | 115 |
| **21** | **44** | 42 | 63 | **63** | **88** | 84 | 117 |
| **22** | **45** | 43 | 64 | **64** | **89** | 85 | 118 |
| **23** | **46** | 44 | 66 | **65** | **90** | 86 | 120 |
| **24** | **47** | 45 | 67 | **66** | **91** | 87 | 121 |
| **25** | **47** | 46 | 68 | **67** | **93** | 88 | 123 |
| **26** | **48** | 47 | 69 | **68** | **94** | 89 | 124 |
| **27** | **49** | 48 | 70 | **69** | **95** | 90 | 126 |
| **28** | **50** | 49 | 71 | **70** | **97** | 91 | 127 |
| **29** | **51** | 50 | 72 | **71** | **98** | 92 | 129 |
| **30** | **52** | 51 | 73 | **72** | **99** | 93 | 131 |
| **31** | **53** | 52 | 74 | **73** | **101** | 94 | 132 |
| **32** | **54** | 53 | 75 | **74** | **102** | 95 | 134 |
| **33** | **55** | 54 | 77 | **75** | **104** | 96 | 135 |
| **34** | **56** | 55 | 78 | **76** | **105** | 97 | 137 |
| **35** | **56** | 56 | 79 | **77** | **106** | 98 | 139 |
| **36** | **57** | 57 | 80 | **78** | **108** | 99 | 140 |
| **37** | **58** | 58 | 81 | **79** | **109** | 100 | 142 |
| **38** | **59** | 59 | 83 | **80** | **111** |  |  |
| **39** | **60** | 60 | 84 | **81** | **112** |  |  |
| **40** | **61** | 61 | 85 | **82** | **114** |  |  |

**Table 5.** CCI informant-assessment (20 items) mapped to ECog informant-assessment (39 items) using Quadratic Regression

| **CCI-20** | **ECog** | **CCI-20** | **ECog** | **CCI-20** | **ECog** | **CCI-20** | **ECog** |
| --- | --- | --- | --- | --- | --- | --- | --- |
| **20** | **42** | 41 | 63 | **62** | **91** | 83 | 125 |
| **21** | **43** | 42 | 65 | **63** | **93** | 84 | 127 |
| **22** | **44** | 43 | 66 | **64** | **94** | 85 | 129 |
| **23** | **45** | 44 | 67 | **65** | **96** | 86 | 131 |
| **24** | **45** | 45 | 68 | **66** | **97** | 87 | 132 |
| **25** | **46** | 46 | 70 | **67** | **99** | 88 | 134 |
| **26** | **47** | 47 | 71 | **68** | **100** | 89 | 136 |
| **27** | **48** | 48 | 72 | **69** | **102** | 90 | 138 |
| **28** | **49** | 49 | 73 | **70** | **103** | 91 | 140 |
| **29** | **50** | 50 | 75 | **71** | **105** | 92 | 142 |
| **30** | **51** | 51 | 76 | **72** | **107** | 93 | 144 |
| **31** | **52** | 52 | 77 | **73** | **108** | 94 | 145 |
| **32** | **53** | 53 | 79 | **74** | **110** | 95 | 147 |
| **33** | **55** | 54 | 80 | **75** | **112** | 96 | 149 |
| **34** | **56** | 55 | 81 | **76** | **113** | 97 | 151 |
| **35** | **57** | 56 | 83 | **77** | **115** | 98 | 153 |
| **36** | **58** | 57 | 84 | **78** | **117** | 99 | 155 |
| **37** | **59** | 58 | 85 | **79** | **118** | 100 | 156 |
| **38** | **60** | 59 | 87 | **80** | **120** |  |  |
| **39** | **61** | 60 | 88 | **81** | **122** |  |  |
| **40** | **62** | 61 | 90 | **82** | **123** |  |  |

**Table 6.** CCI-12 (12 items) self-assessment mapped to ECog self-assessment using Quadratic Regression

| **CCI-12** | **ECog** | **CCI-12** | **ECog** | **CCI-20** | **ECog** |
| --- | --- | --- | --- | --- | --- |
| **12** | **37** | 33 | 55 | **54** | **77** |
| **13** | **38** | 34 | 56 | **55** | **78** |
| **14** | **39** | 35 | 56 | **56** | **79** |
| **15** | **40** | 36 | 57 | **57** | **80** |
| **16** | **40** | 37 | 58 | **58** | **81** |
| **17** | **41** | 38 | 59 | **59** | **83** |
| **18** | **42** | 39 | 60 | **60** | **84** |
| **19** | **43** | 40 | 61 |  |  |
| **20** | **43** | 41 | 62 |  |  |
| **21** | **44** | 42 | 63 |  |  |
| **22** | **45** | 43 | 64 |  |  |
| **23** | **46** | 44 | 66 |  |  |
| **24** | **47** | 45 | 67 |  |  |
| **25** | **47** | 46 | 68 |  |  |
| **26** | **48** | 47 | 69 |  |  |
| **27** | **49** | 48 | 70 |  |  |
| **28** | **50** | 49 | 71 |  |  |
| **29** | **51** | 50 | 72 |  |  |
| **30** | **52** | 51 | 73 |  |  |
| **31** | **53** | 52 | 74 |  |  |
| **32** | **54** | 53 | 75 |  |  |

**Table 7.** CCI-12 (12 items) informant-assessment mapped to ECog informant-assessment using Quadratic Regression

| **CCI-12** | **ECog** | **CCI-12** | **ECog** | **CCI-20** | **ECog** |
| --- | --- | --- | --- | --- | --- |
| **12** | **35** | 33 | 55 | **54** | **80** |
| **13** | **36** | 34 | 56 | **55** | **81** |
| **14** | **37** | 35 | 57 | **56** | **83** |
| **15** | **38** | 36 | 58 | **57** | **84** |
| **16** | **38** | 37 | 59 | **58** | **85** |
| **17** | **39** | 38 | 60 | **59** | **87** |
| **18** | **40** | 39 | 61 | **60** | **88** |
| **19** | **41** | 40 | 62 |  |  |
| **20** | **42** | 41 | 63 |  |  |
| **21** | **43** | 42 | 65 |  |  |
| **22** | **44** | 43 | 66 |  |  |
| **23** | **45** | 44 | 67 |  |  |
| **24** | **45** | 45 | 68 |  |  |
| **25** | **46** | 46 | 70 |  |  |
| **26** | **47** | 47 | 71 |  |  |
| **27** | **48** | 48 | 72 |  |  |
| **28** | **49** | 49 | 73 |  |  |
| **29** | **50** | 50 | 75 |  |  |
| **30** | **51** | 51 | 76 |  |  |
| **31** | **52** | 52 | 77 |  |  |
| **32** | **53** | 53 | 79 |  |  |

## DISCUSSION

The purpose of this study was to examine the relationship between the CCI and the ECog to facilitate future harmonized cross-study and longitudinal assessments of perceived cognitive change. We found high correlations between the CCI and ECog in self and informant reports across all overlapping domains, which supported the development of a co-calibrated crosswalk to translate between scores on the two assessments. The highest correlations for self and informant assessments were between the ECog and CCI total scores. Due to the content differences between the instruments, which in the ECog includes visuospatial questions and the CCI does not, the high correlations despite these differences suggest that the visual-spatial items in the ECog do not have a large impact on the total score.

. Histograms of all the self-report CCI and ECog and the informant-report CCI and ECog total scores were plotted to assess frequency distributions (Figure 1). While the highest number of entries in the informant report histogram included the minimum total score, the self report histogram did not. This could suggest that participants more often report mild cognitive concerns compared to informants. However, there are limitations to this conclusion because different participants were included in the self and informant cohorts. Previous research has shown that informant reports are more strongly correlated to biomarkers of neurodegenerative disease [15]. Two tables were made to translate between the ECog and CCI: one for self assessments and one for informant assessments. We were able to map the CCI total score to an equivalent ECog total score using a quadratic equation. In the self-evaluation data, a total score of 20 on the CCI did not map below 43 on the ECog, even though the minimum score is 39. A score of 100 on the CCI did not map above a score of 142 on the ECog, although the maximum score is 156 (Table 4). In the informant-evaluation data, a score of 20 did not map below 42 and a score of 100 mapped to 157. For the purposes of the translation table, the maximum ECog score was capped at 156 (Table 5).

Limitations of this study include generalizability, due to the high mean education and limited portion of the sample from underrepresented groups. Other studies using these instruments are currently working on replication and extension in more diverse and representative community-based samples. Additionally, the CCI and ECog were not given at precisely the same time, which may have impacted our results. Despite these limitations, co-calibration and translation between scales is feasible and of practical value.

In summary, this study contributes analysis and harmonization methodology for two commonly used instruments to assess self-perceived and proxy-based cognitive concerns. This is important to foster additional research on varying AD prodromal stages, in which SCD concerns are among the first reported symptoms. Total scores of the CCI and ECog assessments show strong correlations and validate the harmonization of these tools to assess longitudinal change in cognitive concerns. Future longitudinal research is needed to characterize the relationship between these assessments and long-term clinical outcomes and biomarker trajectories. Self and informant reports should also be analyzed for other neurodegenerative diseases to better understand potential implications for differential diagnosis, longitudinal follow-up, and other clinical applications.

## Data Availability

Data used in the preparation of this article were obtained from the Alzheimer's Disease Neuroimaging Initiative (ADNI; http://adni.loni.usc.edu) and the Indiana Alzheimer's Disease Research Center (IADRC) including participants from the Indiana Aging and Memory Study (IMAS) initial phase and IADRC phases.

## ACKNOWLEDGEMENTS

This research was supported by grants from the Indiana CTSI (UL1 TR002529) and the National Institute on Aging (U01 AG024904, R01 AG19771, P30 AG010133, P30 AG072976, K01 AG049050, R01 AG067188, U01 AG068057 and U01 AG072177).

Data collection and sharing for this project was funded by the Alzheimer’s Disease Neuroimaging Initiative (ADNI) (National Institutes of Health Grant U01 AG024904) and DOD ADNI (Department of Defense award number W81XWH-12-2-0012). See the ADNI website (http://adni.loni.usc.edu) for more details.

